# Distress among Brazilian university students due to the Covid-19 pandemic: survey results and reflections

**DOI:** 10.1101/2020.06.19.20135251

**Authors:** Carlos Von Krakauer Hübner, Marcella de Lima Bruscatto, Rafaella Dourado Lima

**Author notes:** **Corresponding author** Carlos von Krakauer Hübner PhD, Washington Luiz Ave, 831, Sorocaba, SP, 18031-000, Brazil. **Contributor and guarantor information** All authors have contributed to the planning, conduct and reporting of the work described in the article. The corresponding author attests that all listed authors meet authorship criteria and that no others meeting the criteria have been omitted, and affirms that this manuscript is an honest, accurate, and transparent account of the study being reported; that no important aspects of the study have been omitted; and that any discrepancies from the study as planned (and, if relevant, registered) have been explained.

## Abstract

The first case of infection with the new coronavirus was identified in December 2019 in Wuhan, China. In March, the World Health Organization (WHO) defined the disease epidemic as a pandemic. Thus, a quarantine was imposed by many governments. As a consequence, and given that epidemiological outbreaks of infectious diseases, such as Covid-19, are associated with psychological disorders and symptoms of mental illness, researchers at the Shanghai Mental Health Center have created the Covid-19 Peritraumatic Distress Index (CPDI), in which the results are obtained: normal, mild/moderate distress and severe distress. The main objective of the study was based on the application of CPDI, in order to identify the health and well-being of Brazilian students from different undergraduate courses at the Pontifical Catholic University of São Paulo (PUC/SP) during the Covid-19 pandemic and to test the hypothesis that medical students suffer more than students from other courses. The research is based on a cross-sectional observational study, in which we applied, using Google Forms^R^, the questions contained in CPDI, among with demographic data: age, sex, educational institution, undergraduate course and school year. The Index was applied online for seven days in which a total of 654 valid responses were obtained: 501 (76.6%) female and 149 (22.8%) male. Regarding age, 333 students (50.91%) were 17-20 years old, 279 (42.66%) between 21-25, 30 (4.59%) between 26-30 and 12 (1.84%) between 31-50. The results indicate that the participants reported significant psychological distress, according to the CPDI score. Practically 90% (87.92%) of the students experienced suffering, while only 12.08% did not suffer. The study provides the first empirical evidence on the level of psychological distress in Brazilian university students during the Covid-19 pandemic. Also, it suggests support and monitoring of university students during and after the pandemic, with effective and efficient intervention in their mental health.

**Summary boxes:** *SECTION 1:* Researchers at the Shanghai Mental Health Center have created the Covid-19 Peritraumatic Distress Index (CPDI), to measure the amount of psychological suffering of the population, due to the pandemic of Coronavirus. Before our research, the survey had been applicated in China and Iran. The main objective of the study was to identify with the survey, the health and well-being of Brazilian students from different undergraduate courses at the Pontifical Catholic University of São Paulo (PUC/SP) during the Covid-19 pandemic and to test the hypothesis that medical students suffer more than students from other courses.

*SECTION 2:* The study provides the first empirical evidence on the level of psychological distress in Brazilian university students during the Covid-19 pandemic, practically 90% (87.92%) of the students experienced some suffering.

## INTRODUTION

The first case of infection by the new coronavirus was identified in December 2019, in Wuhan, China^1^. Since then, cases have spread rapidly across the world. In February, Brazil’s first case was identified in São Paulo. At April 24, 2020, in the country, there were 52,995 confirmed cases and 3670 deaths, while in the world the number of cases totals 2.79 million and a total of 196 thousand deaths^2^. In March, the World Health Organization (WHO) defined the disease epidemic as a progressive pandemic.^3^

Covid-19, the name given to the disease caused by the new coronavirus, SARS-CoV2^4^, despite not having its pathophysiology completely elucidated, presents with a clinical picture of pneumonia, fever, difficulty breathing and pulmonary infection^5,6^, associated with a disseminated intravascular coagulation (DIC) ^7,8^. As it is a disease whose transmissibility occurs through one sick person to another or through close contact^9^, the best form of prevention is social isolation^10^. Therefore, quarantine was established by many governments. As a consequence, and given that epidemiological outbreaks of infectious diseases, such as Covid-19, are associated with psychological distress and symptoms of mental illness^11–13^, many people have their lives impacted and this should have an important impact on their health, since in 1947 the WHO developed the concept of health as “a state of complete physical, mental and social well-being and not just the absence of disease or infirmity” ^14^. For instance, it is worth mentioning that medical students in China, during the pandemic, experienced more stress and anxiety.^15^

Researchers at the Shanghai Mental Health Center, during the epidemic, developed a questionnaire to research and assess the level of peritraumatic distress specific to Covid-19. The word distress represents an act or effect of suffering, physical pain, anguish, affliction, bitterness, patience and resignation^16^. The questionnaire, called the Covid-19 Peritraumatic Distress Index (CPDI), has a ranking from 0 to 100. A score between 28 and 51 indicates mild to moderate psychological distress. A score ≥ 52 indicates severe psychological distress. CPDI inquires about the frequency of anxiety, depression, specific phobias, cognitive change, avoidance and compulsive behavior, obsessive thoughts, physical symptoms and loss of social functioning. The questionnaire’s reliability criterion, verified by the researchers, is based on Cronbach’s alpha coefficient, which, in this case, is 0.95 (p <0.001).^17^

The main objective of the study was based on the application of the Covid-19 Peritraumatic Distress Index, in order to identify the health and well-being of Brazilian students from different undergraduate courses at the Pontifical Catholic University of São Paulo (PUC/SP) during the Covid-19 pandemic and to test the hypothesis that medical students suffer more than students from other courses. Besides that, quantify the prevalence and severity of psychological distress in the sample studied, and therefore provide data to adapt and implement relevant mental health intervention policies to deal with this challenge in an efficient and effective manner, prioritizing assistance to psychologically affected people.

## METHODS

The survey was approved by the ethic committee of Pontifical Catholic University of São Paulo, was voluntary, and we assured the participants confidentiality and anonymity of their responses, evidenced by the free and informed consent term (can be seen in the appendix), in which the participants had to read and accept it in order to answer it.

The inclusion criteria used were undergraduate students, specifically from the Pontifical Catholic University of São Paulo, using the electronic form. Those who were not undergraduate students enrolled at the Pontifical Catholic University of São Paulo were excluded, as well as those who, perhaps, were not actively enrolled.

The study uses a cross-sectional observational analytical primary study design, evaluating medical students and other undergraduate courses at PUC/SP. To them it was sent a semi-structured and self-administered questionnaire, answered electronically, using Google Forms^R^, with the questions contained in CPDI along with demographic data: age, gender, educational institution, undergraduate course and the respective academic year.

The Questionnaire contains 24 questions, each of which has 5 answers, whose score is 0 to 4 points for each question. At the end of the questionnaire, the result is obtained from the sum of the answers in addition to the sum of 4 more points. Finally, the points of the responses of each participant, individually, will be added to reach a score. Thus, the compiled data will be analyzed according to the following statistics:

1. Description of the distribution of absolute and relative frequencies of the variable degrees of suffering, age, gender, course and grade;
2. Description of the mean and standard deviation of the suffering score variable.
3. Analysis of differences in suffering scores according to age, gender, course and grade, with special attention to the medical school and its boarding school, using linear regression.

For all tests, a significance level of 0.05, bilateral, were adopted.

The English and Portuguese versions of CPDI, as well as its scoring key can be found in the appendix.

## RESULTS

The COVID-19 Peritraumatic Distress Index (CPDI) was applied online for seven days, from 12 May 2020 to 18 May 2020, in which a total of 764 responses were obtained. Of these, 110 were excluded, as these participants did not belong to the group of students at the Pontifical Catholic University of São Paulo (PUC/SP), a mandatory inclusion criterion. Thus, the total of valid responses analyzed was 654, among them: 501 (76.6%) female, 149 (22.8%) male and 4 (0.6%) preferred not to say the gender. Regarding age, 333 students (50.91%) were between 17 and 20 years old, 279 (42.66%) between 21 and 25 years old, 30 (4.59%) between 26 and 30 years old and 12 (1.84%) between 31 and 50 years old. Concerning the scores, 87.92% of the entire sample presented some level of psychological distress (52.90% mild/moderate and 35.02% severe). Regarding graduation, responses were obtained from 27 different courses (Table 1).

**Table 1.** Relative and Absolute frequency of undergraduate courses at the Pontifical Catholic University of São Paulo and their respective academic years.

|  | 1st year | 2nd year | 3rd year | 4th year | 5th year | 6th year | Total |
| --- | --- | --- | --- | --- | --- | --- | --- |
| **Computer Science** | 8 (1,224%) | 1 (0,153%) | 1 (0,153%) | 0 (0%) | 0 (0%) | 0 (0%) | **10 (1,53%)** |
| **Medicine** | 17 (2,6%) | 10 (1,53%) | 14 (2,141%) | 10 (1,53%) | 26 (3,976%) | 42 (6,423%) | **119 (18,196%)** |
| **International Relations** | 24 (3,67%) | 13 (1,988%) | 9 (1,377%) | 8 (1,224%) | 0 (0%) | 0 (0%) | **54 (8,257%)** |
| **Psychology** | 6 (0,918%) | 8 (1,224%) | 13 (1,988%) | 5 (0,765%) | 4 (0,612%) | 0 (0%) | **36 (5,505%)** |
| **Administration** | 14 (2,141%) | 12 (1,835%) | 8 (1,224%) | 3 (0,459%) | 4 (0,612%) | 0 (0%) | **41 (6,27%)** |
| **Pedagogy** | 12 (1,835%) | 6 (0,918%) | 7 (1,071%) | 4 (0,612%) | 0 (0%) | 0 (0%) | **29 (4,435%)** |
| **Physiotherapy** | 12 (1,835%) | 8 (1,224%) | 14 (2,141%) | 7 (1,071%) | 5 (0,765%) | 0 (0%) | **46 (7,034%)** |
| **Accounting Sciences** | 3 (0,459%) | 1 (0,153%) | 0 (0%) | 2 (0,306%) | 0 (0%) | 0 (0%) | **6 (0,918%)** |
| **Biomedical Engineering** | 6 (0,918%) | 1 (0,153%) | 2 (0,306%) | 1 (0,153%) | 1 (0,153%) | 0 (0%) | **11 (1,682%)** |
| **Nursing** | 8 (1,224%) | 9 (1,377%) | 4 (0,612%) | 6 (0,918%) | 0 (0%) | 0 (0%) | **27 (4,129%)** |
| **Economic Sciences** | 18 (2,754%) | 9 (1,377%) | 12 (1,836%) | 5 (0,765%) | 2 (0,306%) | 0 (0%) | **46 (7,035%)** |
| **Design** | 2 (0,306%) | 5 (0,765%) | 6 (0,918%) | 0 (0%) | 0 (0%) | 0 (0%) | **13 (1,988%)** |
| **Law** | 11 (1,682%) | 31 (4,741%) | 4 (0,612%) | 7 (1,071%) | 21 (3,212%) | 0 (0%) | **74 (11,315%)** |
| **Jornalism** | 16 (2,447%) | 2 (0,306%) | 14 (2,141%) | 14 (2,141%) | 1 (0,153%) | 0 (0%) | **47 (7,187%)** |
| **Languages** | 0 (0%) | 6 (0,918%) | 0 (0%) | 0 (0%) | 0 (0%) | 0 (0%) | **6 (0,918%)** |
| **Social Service** | 13 (1,988%) | 0 (0%) | 0 (0%) | 0 (0%) | 1 (0,153%) | 0 (0%) | **14 (2,141%)** |
| **Theology** | 0 (0%) | 0 (0%) | 0 (0%) | 2 (0,306%) | 0 (0%) | 0 (0%) | **2 (0,306%)** |
| **Actuarial Science** | 2 (0,306%) | 3 (0,459%) | 4 (0,612%) | 1 (0,153%) | 0 (0%) | 0 (0%) | **10 (1,53%)** |
| **Marketing And Advertising** | 12 (1,835%) | 3 (0,459%) | 2 (0,306%) | 0 (0%) | 0 (0%) | 0 (0%) | **17 (2,6%)** |
| **Body Arts Communication** | 4 (0,612%) | 0 (0%) | 2 (0,306%) | 0 (0%) | 1 (0,153%) | 1 (0,153%) | **8 (1,224%)** |
| **Production Engineering** | 0 (0%) | 2 (0,306%) | 1 (0,153%) | 1 (0,153%) | 3 (0,459%) | 0 (0%) | **7 (1,071%)** |
| **Speech-Language Pathology** | 5 (0,765%) | 10 (1,53%) | 0 (0%) | 1 (0,153%) | 0 (0%) | 0 (0%) | **16 (2,447%)** |
| **Philosophy** | 1 (0,153%) | 1 (0,153%) | 0 (0%) | 0 (0%) | 0 (0%) | 0 (0%) | **2 (0,306%)** |
| **Social Sciences** | 2 (0,306%) | 1 (0,153%) | 1 (0,153%) | 0 (0%) | 0 (0%) | 0 (0%) | **4 (0,612%)** |
| **History** | 4 (0,612%) | 2 (0,306%) | 0 (0%) | 1 (0,153%) | 0 (0%) | 0 (0%) | **7 (1,071%)** |
| **Communication and Multimedia** | 0 (0%) | 0 (0%) | 1 (0,153%) | 0 (0%) | 0 (0%) | 0 (0%) | **1 (0,153%)** |
| **Data Science and Artificial Intelligence** | 1 (0,153%) | 0 (0%) | 0 (0%) | 0 (0%) | 0 (0%) | 0 (0%) | **1 (0,153%)** |
| **Total** | **201 (30,734%)** | **144 (22,019%)** | **119 (18,196%)** | **78 (11,927%)** | **69 (10,551%)** | **43 (6,575%)** | **654 (100%)** |

## DISCUSSION

The results indicate that the participants reported significant psychological distress, according to the CPDI score. Practically 90% (87.92%) of the students experienced suffering (52.90% of the participants with scores between 28 and 51, and 35.02% with scores ≥ 52), while only 12.08% did not suffer, obtaining a mean of 45.70, with a standard deviation (SD) of 14.88, in a total sample of 654. The results are alarming, especially when compared to those of the Chinese and Iranian studies, whose means were 23.65 (SD: 5.45) and 34.54 (SD: 14.92), respectively.

It is important to point out that when the research was started the state of São Paulo, where the University is located, in Brazil, was already in its 50th day of quarantine, and according to the Ministry of Health, it already had 47.719 cases infected by Covid-19 in the state, in addition to 177.589 confirmed cases and 12.400 deaths in the country.^18^ Based on these data, it is possible to speculate a correlation between the increase in psychological distress and these stressors.

Through statistical analysis of multiple linear regression, a high rate of peri traumatic distress was associated with the female gender, with undergraduate courses that not Medicine and with non-medical interns. The women who responded showed higher psychological distress when compared to men (mean (SD) = 47.14 (14.42) vs 40.83 (15.41), *p =* 0.001, 95% *CI* 1,26 to 2,50), which enhances the results from previous research which concluded that women are much more vulnerable to stress and more likely to develop post-traumatic stress.^19^

Medical interns reported a lower score of suffering compared to non-intern students (mean (SD) = 50.12 (13.11) vs 38.04 (14.02), *p =* 0.001, 95% *CI* 3,16 to 4,14) and to students from other courses (mean (SD) = 42.20 (14.82) vs 46.48 (14.79), *p =* 0.004, 95% *CI* 1,26 to 2,69). This suggests that the level of health knowledge, specifically the medical one, is a protective factor.

Age did not appear to be a significant predictor of psychological distress (p=0.738), as occurred in a Chinese study sample.^20^

The study has certain limitations. First, the research is based on a cross-sectional observational study. Therefore, there was no follow-up of individuals. Second, our sample does not reflect the national college students, since the objective was to evaluate the students from PUC/SP. Third, although there were responses from several undergraduate courses at the University, some of them presented a very small sample number, such as Communication and Multimedia (0.153%), Data Science and Artificial Intelligence (0.153%), Theology (0.306%), Philosophy (0.306%), when compared to other courses, such as Medicine (18.196%).

The study provides the first empirical evidence on the level of psychological distress in Brazilian university students during the Covid-19 pandemic. The results demonstrate a significant psychological disorder in the participants. Therefore, the study suggests support and monitoring of university students during and after the pandemic, with effective and efficient intervention in their mental health, such as (1) greater attention to vulnerable groups, such as women, non-medical interns and students of others courses that not Medicine; (2) University strategy, with planning and coordination for psychological and medical care and assistance for psychologically affected students, in an accessible and free way; (3) future monitoring, in order to ensure longitudinal monitoring of those with psychological distress, as well as an active search for new cases; (4) joint efforts with other universities, expanding the screening of students with some level of mental distress or mental anguish.

## Data Availability

Google Drive created with the objective of making available all data belonging to the manuscript "Distress among Brazilian university students due to the Covid-19 pandemic: survey results and reflections".
https://drive.google.com/drive/folders/1YJWCo1HydFYvAEyFcAo7rv15z46sZLwr?usp=sharing

## Appendix A The Covid-19 Peritraumatic Distress Index (CPDI) in English

Appendix A - The Covid-19 Peritraumatic Distress Index (CPDI) in English
|  | Never | Occasionally | Sometimes | Often | Most of the time |
| --- | --- | --- | --- | --- | --- |
| 1. Compared to usual, I feel more nervous and anxious | 0 | 1 | 2 | 3 | 4 |
| 2. I feel insecure and bought a lot of goods, such as medications, sanitizer, gloves, masks, and/or other home supplies | 0 | 1 | 2 | 3 | 4 |
| 3. I can't stop myself from imagining myself or my family being infected and feel terrified and anxious about it | 0 | 1 | 2 | 3 | 4 |
| 4. I feel empty and helpless no matter what I do | 0 | 1 | 2 | 3 | 4 |
| 5. I feel sympathetic to the Covid-19 patients and their families. I feel sad about them | 0 | 1 | 2 | 3 | 4 |
| 6. I feel helpless and angry about people around me, such as the governors and media | 0 | 1 | 2 | 3 | 4 |
| 7. I am losing faith in the people around me | 0 | 1 | 2 | 3 | 4 |
| 8. I collect information about Covid-19 all day. Even if it's not necessary, I can't stop myself | 0 | 1 | 2 | 3 | 4 |
| 9. I will believe the Covid-19 information from all sources without any evaluation | 0 | 1 | 2 | 3 | 4 |
| 10. I would rather believe in negative news about Covid-19 and be skeptical about the good news | 0 | 1 | 2 | 3 | 4 |
| 11. I am constantly sharing news about Covid-19 (mostly negative news) | 0 | 1 | 2 | 3 | 4 |
| 12. I avoid watching Covid-19 news, since I am too scared to do so | 0 | 1 | 2 | 3 | 4 |
| 13. I am more irritable and have frequent conflicts with my family | 0 | 1 | 2 | 3 | 4 |
| 14. I feel tired and sometimes even exhausted | 0 | 1 | 2 | 3 | 4 |
| 15. Due to feelings of anxiety, my reactions are becoming sluggish | 0 | 1 | 2 | 3 | 4 |
| 16. I find it hard to concentrate | 0 | 1 | 2 | 3 | 4 |
| 17. I find it hard to make any decisions | 0 | 1 | 2 | 3 | 4 |
| 18. During this Covid-19 outbreak, I often feel dizzy, have back pain, or chest discomfort | 0 | 1 | 2 | 3 | 4 |
| 19. During this Covid-19 outbreak, I often feel stomach pain, bloating, or other stomach discomforts | 0 | 1 | 2 | 3 | 4 |
| 20. I feel uncomfortable when communicating with others | 0 | 1 | 2 | 3 | 4 |
| 21. Recently, I rarely talk to my family | 0 | 1 | 2 | 3 | 4 |
| 22. I cannot sleep well. I always dream about myself or my family being infected by coronavirus | 0 | 1 | 2 | 3 | 4 |
| 23. I lost my appetite | 0 | 1 | 2 | 3 | 4 |
| 24. I have constipation or frequent urination | 0 | 1 | 2 | 3 | 4 |

**Note:** CPDI = sum of the score of each question +4.
CPDI ≥ 52: severe distress; CPDI within 29-51: mild/moderate distress; CPDI< 28: Normal.

## Appendix B. The Covid-19 Peritraumatic Distress Index (CPDI) in Portuguese

|  | Nunca | Ocasionalmente | Às vezes | Frequentemente/<br>Muitas vezes | Maior parte do tempo |
| --- | --- | --- | --- | --- | --- |
| 1. Comparado ao habitual, eu me sinto mais nervoso(a) e ansioso(a) | 0 | 1 | 2 | 3 | 4 |
| 2. Eu me sinto inseguro(a) e comprei muitas mercadorias, como medicamentos, desinfetante, luvas, máscaras e/ou outros suprimentos para casa. | 0 | 1 | 2 | 3 | 4 |
| 3. Eu não consigo parar de pensar que eu ou a minha família seja infectada e eu me sinto apavorado(a) e ansioso(a) sobre isso. | 0 | 1 | 2 | 3 | 4 |
| 4. Eu me sinto vazio(a) e impotente não importa o que eu faça. | 0 | 1 | 2 | 3 | 4 |
| 5. Eu sinto compaixão para com os pacientes infectados por Covid-19 e seus familiares. Eu me sinto triste por eles. | 0 | 1 | 2 | 3 | 4 |
| 6. Eu me sinto impotente e bravo(a) com as pessoas à minha volta, assim como de governadores e da mídia. | 0 | 1 | 2 | 3 | 4 |
| 7. Eu estou perdendo a fé nas pessoas à minha volta. | 0 | 1 | 2 | 3 | 4 |
| 8. Eu me informo sobre a Covid-19 o dia todo. Mesmo que não seja necessário, eu não consigo parar. | 0 | 1 | 2 | 3 | 4 |
| 9. Eu acreditarei nas informações sobre Covid-19 de todas as fontes, sem nenhuma avaliação. | 0 | 1 | 2 | 3 | 4 |
| 10. Eu preferiria acreditar em notícias negativas sobre a Covid-19 e ser cético sobre as notícias boas. | 0 | 1 | 2 | 3 | 4 |
| 11. Eu estou constantemente compartilhando notícias sobre Covid-19 (a maioria notícias negativas). | 0 | 1 | 2 | 3 | 4 |
| 12. Eu evito assistir notícias sobre Covid-19, porque estou com medo de fazê-lo. | 0 | 1 | 2 | 3 | 4 |
| 13. Eu estou mais irritado e tenho conflitos frequentes com a minha família. | 0 | 1 | 2 | 3 | 4 |
| 14. Eu me sinto cansado e às vezes até exausto. | 0 | 1 | 2 | 3 | 4 |
| 15. Por causa de sentimentos de ansiedade, minhas reações estão se tornando lerdas. | 0 | 1 | 2 | 3 | 4 |
| 16. Acho difícil me concentrar. | 0 | 1 | 2 | 3 | 4 |
| 17. Acho difícil tomar qualquer decisão. | 0 | 1 | 2 | 3 | 4 |
| 18. Durante esse surto de Covid-19, eu frequentemente sinto tontura, dor nas costas, ou desconforto no peito. | 0 | 1 | 2 | 3 | 4 |
| 19. Durante esse surto de Covid-19 eu frequentemente sinto dor de estômago, inchaço, ou outros desconfortos estomacais. | 0 | 1 | 2 | 3 | 4 |
| 20. Eu me sinto desconfortável quando me comunico com outros. | 0 | 1 | 2 | 3 | 4 |
| 21. Recentemente, eu raramente falo com a minha família. | 0 | 1 | 2 | 3 | 4 |
| 22. Eu não consigo dormir bem. Eu sempre sonho sobre mim ou minha família sendo infectada pelo coronavírus. | 0 | 1 | 2 | 3 | 4 |
| 23. Eu perdi o meu apetite. | 0 | 1 | 2 | 3 | 4 |
| 24. Eu tenho constipação ou urino com frequência. | 0 | 1 | 2 | 3 | 4 |

**Nota** = pontuação elaborada por meio da soma dos pontos + 4
Questionário de Sofrimento Peri traumático de Covid-19 $\geq 52$ : sofrimento psíquico severo; entre 28 – 51: leve/moderado; $< 28$ : normal.

## Appendix C FREE AND CLARIFIED CONSENT

You are being invited as a volunteer to participate in the research “**Distress among Brazilian university students due to the Covid-19 pandemic: survey results and reflections”**.

The first case of infection with the new coronavirus was identified in December 2019 in Wuhan, China. In March, the World Health Organization (WHO) defined the disease epidemic as a progressive pandemic. Thus, the objective of applying the Covid-19 Traumatic Peri Suffering Questionnaire, in order to identify the health and well-being of Brazilian students during the Covid-19 pandemic and to compare the psychological suffering between medical students and medical students other undergraduate courses at the Pontifical Catholic University of São Paulo (PUC-SP). Your anonymity will be guaranteed. The online system, chosen for the application of the questionnaire, does not allow us to identify the respondents to the questionnaire and any material that identifies their participation will be released without their authorization. If you still have doubts, you can be informed about any aspect of the study you want, before or after answering this questionnaire, through the researchers’ contacts.

We clarify that there will be no costs and no payment for those who participate in the study. We consider that there are no risks in participating in this research. At the end of the study, the researchers commit to communicating the results obtained with the research.

This research was analyzed and approved by the Research Ethics Committee of the Pontifical Catholic University of São Paulo, Sorocaba campus. The committee is located at Rua Joubert Wey, 290, Vergueiro, Sorocaba - SP and can receive any complaints or provide other information about the research by calling 15-3212-9896 or, during business hours. Any questions, please contact those responsible for the research:

Prof. Dr. Carlos von Krakauer Hübner: telephone (15) 3232 8933 or by

The students will also participate in the project:

Marcella de Lima Bruscatto: phone (11) 98741-8222

Rafaella Dourado Lima: telephone (11) 99975-4695 or

POST-CLARED CONSENT

() I declare that, after having been properly explained by the researcher and having understood what was explained to me in this term, I agree to participate in the research project. Your acceptance to participate will be considered as your electronic signature of this informed consent form and the questionnaire link will only be available after your acceptance.

